# Sleep-Disordered Breathing Destabilizes Ventricular Repolarization

**DOI:** 10.1101/2023.02.10.23285789

**Authors:** Soroosh Solhjoo, Mark C. Haigney, Trishul Siddharthan, Abigail Koch, Naresh M. Punjabi

**Author notes:** Corresponding Author: Soroosh Solhjoo, PhD, F. Edward Hébert School of Medicine, 4301 Jones Bridge Road, Bethesda, MD 20814.

## Abstract

**Rationale:** Sleep-disordered breathing (SDB) increases the risk of cardiac arrhythmias and sudden cardiac death.

**Objectives:** To characterize the associations between SDB, intermittent hypoxemia, and the beat-to-beat QT variability index (QTVI), a measure of ventricular repolarization lability associated with a higher risk for cardiac arrhythmias, sudden cardiac death, and mortality.

**Methods:** Three distinct cohorts were used for the current study. The first cohort, used for cross-sectional analysis, was a matched sample of 122 participants with and without severe SDB. The second cohort, used for longitudinal analysis, consisted of a matched sample of 52 participants with and without incident SDB. The cross-sectional and longitudinal cohorts were selected from the Sleep Heart Health Study participants. The third cohort comprised 19 healthy adults exposed to acute intermittent hypoxia and ambient air on two separate days. Electrocardiographic measures were calculated from one-lead electrocardiograms.

**Results:** Compared to those without SDB, participants with severe SDB had greater QTVI (-1.19 in participants with severe SDB vs. -1.43 in participants without SDB, *P* = 0.027), heart rate (68.34 vs. 64.92 beats/minute; *P* = 0.028), and hypoxemia burden during sleep as assessed by the total sleep time with oxygen saturation less than 90% (TST_90_; 11.39% vs. 1.32%, *P* < 0.001). TST_90_, but not the frequency of arousals, was a predictor of QTVI. QTVI during sleep was predictive of all-cause mortality. With incident SDB, mean QTVI increased from -1.23 to -0.86 over 5 years (*P* = 0.017). Finally, exposing healthy adults to acute intermittent hypoxia for four hours progressively increased QTVI (from -1.85 at baseline to -1.64 after four hours of intermittent hypoxia; *P* = 0.016).

**Conclusions:** Prevalent and incident SDB are associated with ventricular repolarization instability, which predisposes to ventricular arrhythmias and sudden cardiac death. Intermittent hypoxemia destabilizes ventricular repolarization and may contribute to increased mortality in SDB.

## INTRODUCTION

Sleep-disordered breathing (SDB) is a common disorder affecting 9%-38% of the general population (1, 2). SDB is more prevalent in older adults, affecting approximately 90% of men and 78% of women between the ages of 65 and 80 (3). Recurrent episodes of apneas and hypopneas in SDB are associated with intermittent hypoxemia and frequent arousals from sleep. There is unequivocal evidence that SDB is associated with hypertension, cardiovascular disease, and a higher predisposition for cardiac arrhythmias (4–11). Conduction abnormalities, bradyarrhythmias, and both atrial and ventricular tachyarrhythmias are more prevalent in people with SDB than in those without (11). A growing body of empirical evidence also indicates that SDB is a risk factor for sudden cardiac death (12–15).

The role of SDB in triggering malignant arrhythmias can be assessed through its association with abnormal electrocardiographic (ECG) features associated with a higher risk for sudden cardiac death. It is well established that the beat-to-beat QT variability index (QTVI), a measure of ventricular repolarization lability, is predictive of ventricular arrhythmias and sudden cardiac death (16–21). Elevated QTVI has also been linked to a higher propensity for all-cause and cardiovascular mortality (22). An increase in QTVI indicates that the QT interval is varying out of proportion to heart rate variability. SDB-associated nocturnal intermittent hypoxemia and sleep fragmentation can increase the sympathetic nervous system’s activity, which, in turn, can modulate the QT interval through mechanisms independent of its effect on heart rate (23–25) and result in an increase in QT variability that is out of proportion to the variability in heart rate (26–28). Therefore, it is likely that SDB could increase QTVI. However, the current body of evidence is limited, and significant gaps remain in our understanding of the effects of SDB on QTVI. First, available data are conflicting, with studies showing no association (29), a positive association (30), or an inverse association (31) between SDB severity and QTVI. Second, it remains to be determined whether incident SDB leads to longitudinal changes in ventricular repolarization. Finally, there are no empirical data as to whether intermittent hypoxemia, a pathophysiological concomitant of SDB, can independently alter QTVI. Therefore, the current study sought to determine whether prevalent and incident SDB are associated with ventricular repolarization instability and to characterize the role of intermittent hypoxemia in increasing QTVI.

## METHODS

### Study Design and Sample Selection

Three distinct cohorts were selected for this study. The first two cohorts were derived from the Sleep Heart Health Study (SHHS), a longitudinal study on the cardiovascular consequences of SDB (32–34). Of the 6,441 participants who completed the baseline SHHS examination and polysomnography, 3,296 had a follow-up polysomnogram approximately five years later. The home polysomnogram was conducted using a portable monitor (P-Series, Compumedics, Charlotte, NC). The following physiologic signals were recorded: C3 and C4 electroencephalograms, right and left electrooculograms, a single-lead ECG (sampled at 125 Hz for baseline and 250 Hz for follow-up), a chin electromyogram, oxyhemoglobin saturation (SpO_2_) by pulse oximetry, chest and abdominal excursion by inductance plethysmography, airflow by an oronasal thermocouple, and body position by a mercury gauge. An apnea was identified if the airflow was absent or nearly absent for at least ten seconds. Hypopnea was defined as at least a 30% reduction in airflow or thoracoabdominal movement below baseline values for at least 10 seconds associated with an oxyhemoglobin desaturation of 4%. Informed consent was obtained from all SHHS participants, and the study protocol was approved by the institutional review committee on human research (Approval Number: NA_00010790). Anonymized data are available from the National Sleep Research Resource (34). To minimize confounding in the current analysis, we excluded participants with prevalent myocardial infarction, coronary artery bypass surgery, heart failure, or angina and those on beta-blockers, calcium channel blockers, or antiarrhythmic agents.

To assess whether prevalent SDB is associated with a higher QTVI, a cross-sectional sample from the baseline SHHS visit was selected that included participants with severe SDB (apnea/hypopnea index (AHI) ≥ 33 events/hour, i.e., 95^th^ percentile) along with a group of age (± 1 year), sex, body mass index (BMI, ± 1 kg/m^2^), and race-matched participants without SDB (AHI < 1.33 events/hour; i.e., 25^th^ percentile). A second sample of SHHS participants was selected to assess whether incident SDB and associated nocturnal hypoxemia increase QTVI over time. The second sample consisted of participants initially without SDB with total sleep time with SpO_2_ less than 90% (TST_90_) < 0.22% (i.e., 50^th^ percentile) who then developed SDB and had a TST_90_ ≥ 10.4% (i.e., 90^th^ percentile) at the follow-up polysomnogram five years later. An age (± 1 year), sex, BMI (± 1 kg/m^2^), and race-matched group of comparator participants without incident SDB was also identified who had a TST_90_ < 0.22% during both the baseline and follow-up polysomnograms.

The decision to use matched samples for the cross-sectional and longitudinal analyses was based on several advantages offered by matching (35). One advantage of matching, particularly pair matching, is that it does not require any assumptions regarding the functional form (i.e., linear vs. nonlinear) of the association between a confounder (e.g., age, sex, prevalent medical conditions) and the outcome (i.e., QTVI). Additionally, matching is advantageous when there are many confounders because it is often challenging to accurately model all the associations between all possible confounders and the outcome, while matching the subgroups is relatively simple. Matching also protects against extrapolation of inferences from the region of covariate overlap between groups. Finally, matching avoids chance associations, which can result from iteratively readjusting a regression model after examining estimates of a particular covariate.

A third cohort of healthy adult volunteers was recruited from the local community as part of an experimental study on the acute effects of intermittent hypoxia, as previously described (36). For each volunteer, the experiment was done on two separate days at the same time in the morning. On one day, the participant was exposed to intermittent hypoxia and, on the other, to normoxia. The order of exposures to intermittent hypoxia or normoxia was random. Intermittent hypoxia was induced using a standard nasal mask connected to a three-way valve by alternating the airflow between hypoxic gas (5% O_2_, 95% N_2_) and ambient air (21% O_2_, 79% N_2_). Exposure to hypoxic gas was maintained until the SpO_2_ reached 85% and was reinstated when SpO_2_ recovered to baseline values. On the normoxia day, the participant received 21% O_2_ for the entire experimental period by replacing the hypoxic gas tank with a pressurized ambient air tank. SpO_2_ was recorded using the Biox 3700 pulse oximeter (Ohmeda, Englewood, CO) at 2 Hz. A one-lead ECG signal was recorded continuously at 200 Hz using an Embla N-7000 system (Natus Medical, Pleasanton, CA). Informed consent was obtained from all participants in the study, and the study protocol was approved by the institutional review committee on human research (Approval Number: NA_00002563).

### ECG Signal Processing

The ECG signal was analyzed using customized software developed in MATLAB (Mathworks, Natick, MA) to derive measures of heart rate (HR), heart rate variability, and QTVI over a predefined epoch length. The standard deviation of the intervals between successive normal beats (SDNN) was calculated as the time-domain measure of heart rate variability. Spectral analysis was used to derive frequency-domain measures of heart rate variability. QT variability was measured using a template-matching algorithm (16, 37) and was calculated as 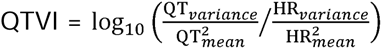. Sensitivity analysis showed no difference between QTVI calculated for ECG signals sampled initially at 250 Hz versus down-sampled to 125 Hz. Therefore, our template-matching QTVI algorithm is robust at the sampling frequencies used for the SHHS cohorts.

### Statistical Analyses

Repeated measurements of electrocardiographic parameters over consecutive epochs were summarized using medians, which were subsequently compared using the *t* test for independent or paired samples as appropriate. Normality was tested using the Lilliefors test. Multivariable linear regression based on least-squares estimation was used to characterize the association between QTVI and hypoxemia burden. The least absolute shrinkage and selection operator (lasso) (38) was used to determine the potential contributors to mortality among the demographic, polysomnographic, and electrocardiographic parameters based on multivariable logistic regression models. Values are reported as the mean ± standard error of the mean (SEM). Statistical analyses and graphing were conducted using the R statistical package, glmnet package (39), MATLAB, and Origin (OriginLab, Northampton, MA).

## RESULTS

### Prevalent SDB and Ventricular Repolarization Instability

Sixty-one participants (45 males, 16 females) with severe SDB met the inclusion criteria of the cross-sectional sample and were matched with 61 participants without prevalent SDB. The average AHI was 46.28 and 0.62 events/hour in participants with and without SDB, respectively. The two groups were matched by age, sex, race, and BMI (Table 1). TST_90_ was greater in those with severe SDB than in those without SDB. The severe-SDB group had a higher mean heart rate and SDNN during sleep than those without SDB. Mean QTVI was also larger in participants with severe SDB, indicating greater ventricular repolarization lability. Stratified analyses by rapid eye movement sleep (REM) and non-REM (NREM) sleep showed similar findings with a larger QTVI with severe SDB irrespective of sleep stage (Figure 1 and Table S1).

**Figure 1.**
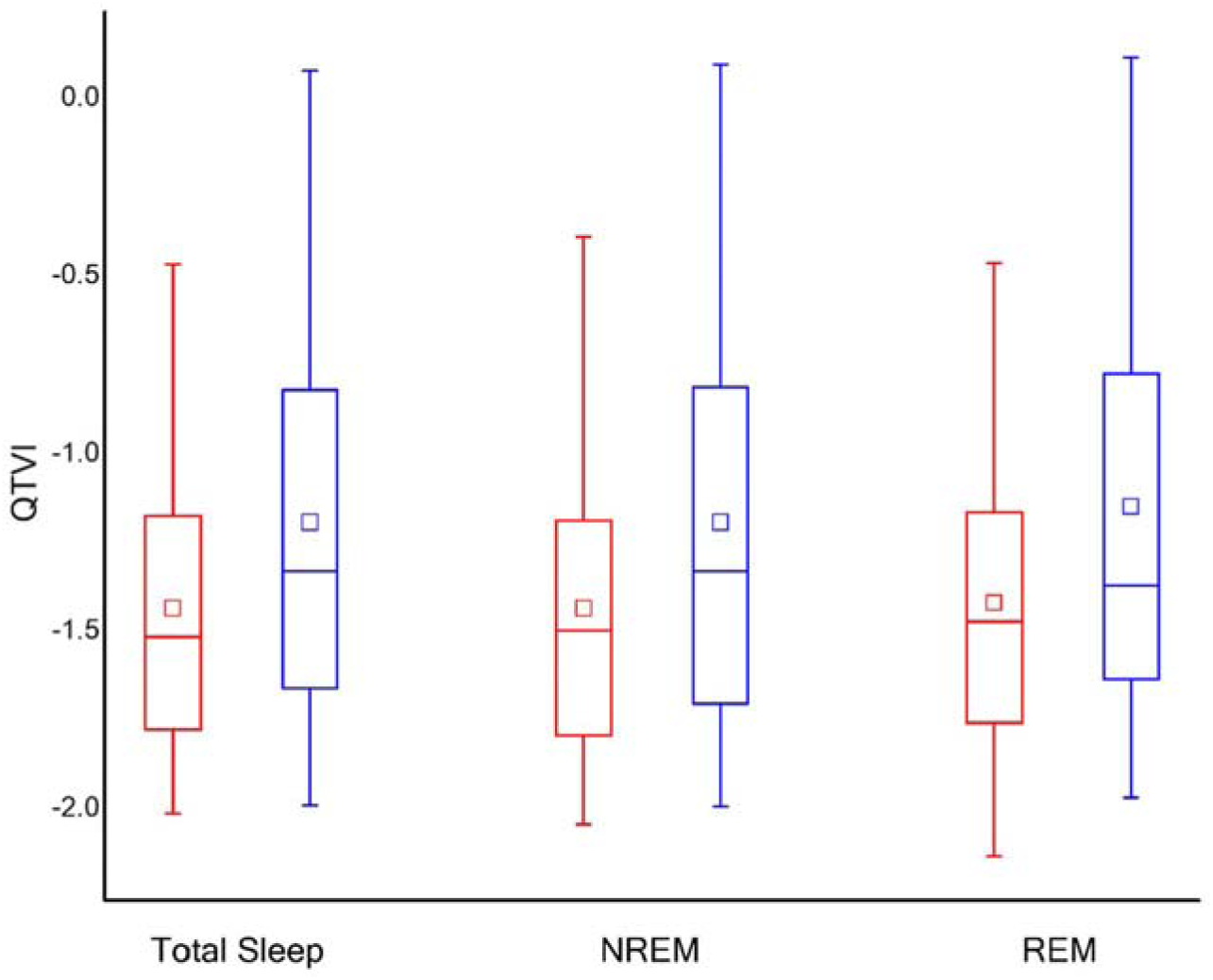
QTVI was higher in participants with severe SDB. The diagram shows the mean QTVI measurements for participants without SDB (N = 61, red) and with severe SDB (N = 61, blue).

**Table 1.** Demographic, polysomnographic, and electrocardiographic data for the cross-sectional sample comparing severe SDB versus no SDB.

| Parameters | No SDB | Severe SDB | *P* value |
| --- | --- | --- | --- |
| Demographic |  |  |  |
| Age (years) | 60.54 (1.00) | 60.72 (1.01) | 0.899 |
| BMI (kg/m<sup>2</sup>) | 29.28 (0.48) | 29.45 (0.50) | 0.802 |
| Polysomnographic |  |  |  |
| AHI (/hour) | 0.62 (0.05) | 46.28 (1.59) | < 0.001 |
| Mean SpO<sub>2</sub> (%) | 95.07 (0.23) | 93.75 (0.20) | < 0.001 |
| TST<sub>90</sub> (%) | 1.32 (0.61) | 11.39 (1.42) | < 0.001 |
| Electrocardiographic |  |  |  |
| QTVI | -1.43 (0.06) | -1.19 (0.08) | 0.027 |
| HR (beats/min) | 64.92 (1.11) | 68.34 (1.08) | 0.028 |
| SDNN (msec) | 32.01 (2.21) | 41.48 (3.25) | 0.018 |
| QTc (msec) | 442.58 (4.80) | 457.86 (4.20) | 0.018 |

Values are the mean (SEM). *P* values are for the Student's *t* test comparing matched participants with and without SDB. N = 61 for each group.

Logistic regression analyses were used to determine whether electrocardiographic (heart rate, SDNN, and QTVI), demographic (age, sex, and BMI), and polysomnographic measures (AHI, frequency of arousals, and TST_90_) were associated with all-cause mortality over an average follow-up of 8.2 years. Heart rate and QTVI were the only two predictors of all-cause mortality selected by the lasso analysis with an area under the receiver operating characteristic curve (AUC) of 0.75 (95% confidence interval: 0.63-0.87) based on 10-fold cross-validation. Including all other covariates in addition to heart rate and QTVI resulted in no significant increase in AUC.

Multivariable linear regression analysis showed that higher TST_90_ values were associated with greater QTVI independent of age, sex, and BMI. For a 10% increase in TST_90_, QTVI increased by 0.13. No association was noted between the frequency of arousals and QTVI.

### Incident SDB and Ventricular Repolarization Instability

Based on the finding that TST_90_ was independently predictive of QTVI in the cross-sectional sample, the role of incident SDB in increasing QTVI was examined. Twenty-six participants (8 males, 18 females) met the criteria for the longitudinal sample with incident SDB and were matched with 26 participants without incident SDB at the 5-year follow-up. Table 2 shows the demographic, polysomnographic, and electrocardiographic data. In participants who developed SDB, AHI increased from 2.54 ± 0.72 events/hour at baseline to 13.74 ± 3.5 events/hour at follow-up. AHI remained unchanged in those without incident SDB. Mean heart rate, SDNN, and QTVI during sleep did not change in those who did not develop SDB between the two visits. However, in those with incident SDB, QTVI increased from -1.23 ± 0.15 to -0.86 ± 0.14 (*P* = 0.017, Figure 2). Stratified analysis by sleep stage revealed that incident SDB was associated with increased QTVI during NREM sleep but not REM sleep (Table S2).

**Figure 2.**
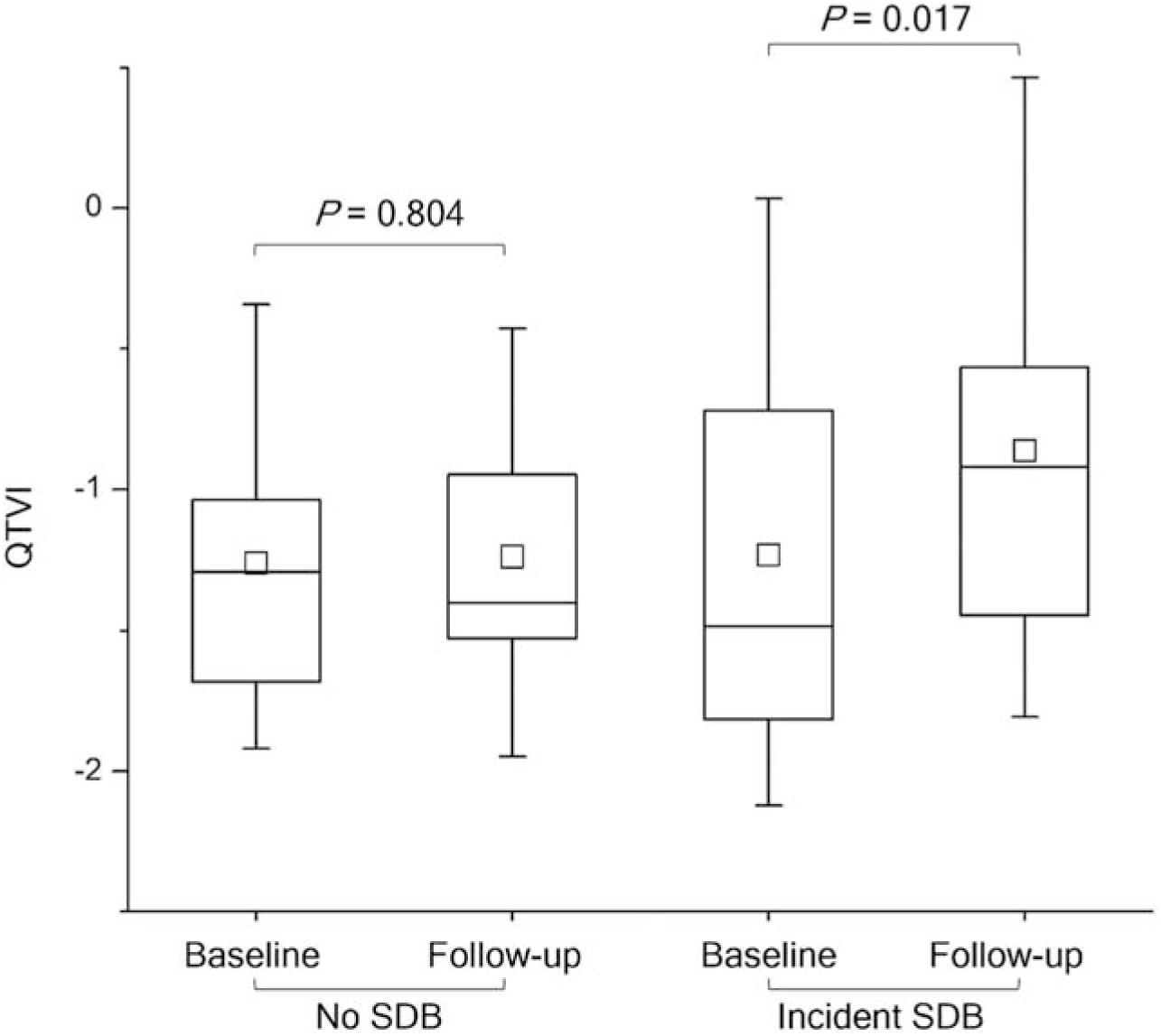
Incident SDB destabilized ventricular repolarization. The diagram shows QTVI for participants with and without incident SDB. QTVI significantly increased in participants who developed SDB. N = 26 in each group. *P* values are for the paired samples *t* test between baseline and follow-up measurements for each group.

**Table 2.** Demographic, polysomnographic, and electrocardiographic data for the longitudinal sample of the participants with and without incident SDB.

| Parameters | Baseline |  |  | Follow-up |  |  |
| --- | --- | --- | --- | --- | --- | --- |
|  | Without SDB | Incident SDB | *P* value | Without SDB | Incident SDB | *P* value |
| **Demographic** |  |  |  |  |  |  |
| Age (years) | 63.88 (1.86) | 64.08 (1.87) | 0.942 | 69.15 (1.83) | 69.12 (1.84) | 0.988 |
| BMI (kg/m<sup>2</sup>) | 27.24 (0.87) | 27.70 (1.00) | 0.732 | 26.87 (0.81) | 28.45 (1.18) | 0.272 |
| **Polysomnographic** |  |  |  |  |  |  |
| AHI (/hour) | 2.79 (0.58) | 2.54 (0.72) | 0.793 | 3.36 (0.67) | 13.74 (3.50) | 0.005 |
| Mean SpO<sub>2</sub> (%) | 95.81 (0.21) | 94.98 (0.24) | 0.012 | 95.59 (0.23) | 91.26 (0.15) | < 0.001 |
| TST<sub>90</sub> (%) | 0.05 (0.01) | 0.05 (0.01) | 0.905 | 0.03 (0.01) | 21.79 (2.08) | < 0.001 |
| **Electrocardiographic** |  |  |  |  |  |  |
| QTVI | -1.26 (0.09) | -1.23 (0.15) | 0.876 | -1.24 (0.10) | -0.86 (0.14) | 0.034 |
| HR (beats/min) | 64.16 (1.51) | 64.68 (1.74) | 0.824 | 64.10 (1.75) | 66.86 (2.08) | 0.315 |
| SDNN (msec) | 28.39 (2.64) | 27.66 (2.69) | 0.846 | 27.43 (2.34) | 24.41 (3.01) | 0.432 |
| QTc (msec) | 436.85 (5.43) | 447.39 (8.40) | 0.297 | 444.93 (5.27) | 450.34 (6.03) | 0.503 |

Values are the mean (SEM). *P* values are for the Student's *t* test comparing participants who developed SDB (N = 26) to those who did not (N = 26).

### Intermittent Hypoxia and Ventricular Repolarization Instability

To assess the adverse effects of acute intermittent hypoxemia on QTVI, ECG data from 19 healthy volunteers exposed to intermittent hypoxia or normoxia were used. The average age and BMI of the 19 healthy volunteers were 25.5 (Range: 18.0–37.0) years and 25.2 (21.7–33.0) kg/m^2^, respectively. On the day of the intermittent hypoxia exposure, the participants were exposed to the hypoxic gas mixture 24.59 ± 0.78 times/hour for an average of 67.52 ± 2.79 seconds per event. Consequently, mean SpO_2_ was significantly lower with intermittent hypoxia (90.90 ± 0.20%) than with normoxia (97.37 ± 0.20%, *P* < 10^-13^, Table S3). Heart rate transiently increased with each episode of hypoxia (Figure 3), leading to increased heart rate variability. SDNN was also higher (*P* = 0.014) during the first hour of intermittent hypoxia than normoxia. Other electrocardiographic measures (i.e., NN interval, LF/(HF+LF), and QTVI) were similar between the two conditions during the first hour. No significant changes were observed in any electrocardiographic measures between the first and the fourth hour of normoxia (Table 3). However, between the first and the last hour of intermittent hypoxia, heart rate and the LF/(LF+HF) ratio increased while log(HF) decreased, indicating a shift in sympathovagal balance towards increased sympathetic activity. QTVI also increased with intermittent hypoxia over the four hours (Table 3). By the 4^th^ hour of exposure, heart rate, SDNN, LF/(LF+HF), and QTVI were significantly greater with intermittent hypoxia than normoxia (Figure 3, right panel).

**Figure 3.**
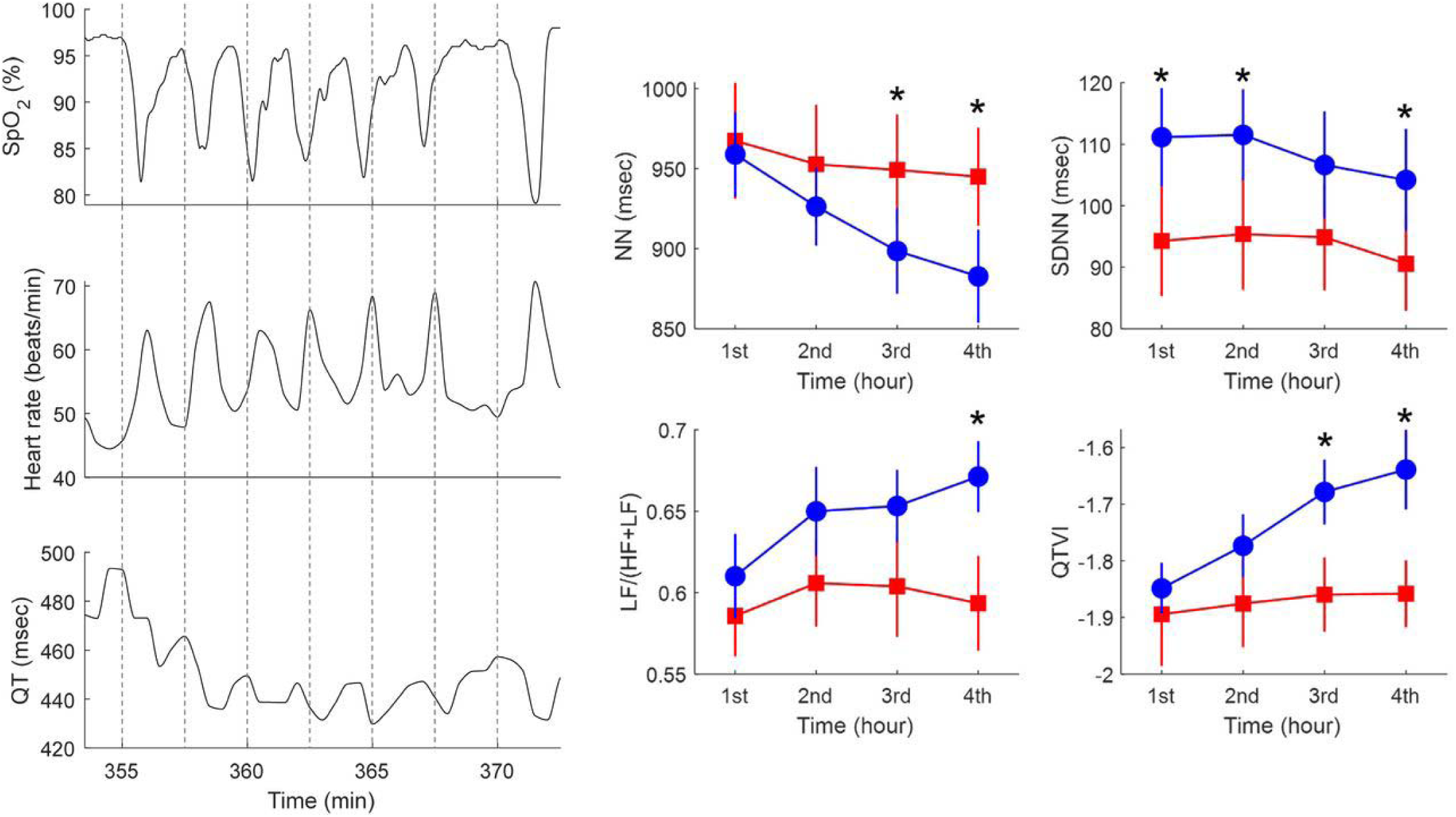
Temporal effects of intermittent hypoxia on electrocardiographic measures. The left panel shows data on one subject during ∼ 20 minutes of exposure to intermittent hypoxia. Cyclical changes were noted in the heart rate and QT interval with intermittent hypoxia. The right panel shows the NN interval, SDNN, LF/(HF+LF) ratio, and QTVI of the 19 subjects for each hour of the four hours of exposure to intermittent hypoxia and normoxia. * indicates *P* < 0.05 for comparing the two conditions during the corresponding hour.

**Table 3.** Electrographic measures during normoxia versus intermittent hypoxia.

| Electrocardiographic Parameters | 1<sup>st</sup> hour | 4<sup>th</sup> hour | *P* value |
| --- | --- | --- | --- |
| Normoxia |  |  |  |
| QTVI | -1.89 (0.09) | -1.86 (0.06) | 0.412 |
| HR (beats/min) | 63.61 (2.43) | 64.73 (2.15) | 0.183 |
| SDNN (msec) | 94.28 (8.99) | 90.56 (7.65) | 0.336 |
| Log(HF) | 3.11 (0.11) | 3.06 (0.10) | 0.189 |
| LF/(HF+LF) | 0.59 (0.02) | 0.59 (0.03) | 0.715 |
| Intermittent hypoxia |  |  |  |
| QTVI | -1.85 (0.04) | -1.64 (0.07) | 0.016 |
| HR | 63.31 (1.51) | 69.21 (2.14) | 0.001 |
| SDNN | 111.15 (7.99) | 104.19 (8.29) | 0.317 |
| Log(HF) | 3.17 (0.09) | 3.01 (0.09) | 0.046 |
| LF/(HF+LF) | 0.61 (0.03) | 0.67 (0.02) | 0.014 |

Values are the mean (SEM). *P* values are for the paired *t* test between normoxia and intermittent hypoxia experiments. N = 19.

## DISCUSSION

The data presented herein demonstrate that SDB is associated with ventricular repolarization instability. In a cohort of middle-aged and older adults, QTVI was larger in participants with severe SDB than in those without SDB. The severity of nocturnal hypoxemia, but not the frequency of arousals, was associated with a larger QTVI, which was, in turn, predictive of all-cause mortality. Longitudinal data showed that incident SDB was associated with increased QTVI over five years. Finally, acute exposure to intermittent hypoxia progressively increased QTVI in a sample of healthy adults.

The last decade has seen enormous growth in the evidence linking SDB to ventricular arrhythmias and sudden cardiac death (12, 14, 40–44). Epidemiological data have shown that SDB is associated with a higher burden of complex ventricular ectopy (i.e., bigeminy, trigeminy, and quadrigeminy), non-sustained ventricular tachycardia, and sudden cardiac death (11). A meta-analysis of 22 studies, which included 42,099 participants, reported that the SDB-related relative risk for sudden death was 1.74 (95% CI: 1.40– 2.10) (44). Subgroup analysis of studies with polysomnography data revealed a dose-response association between SDB severity and the risk of sudden cardiac death (44).

Several biological mechanisms can explicate the observed clinical and epidemiological associations between SDB and sudden cardiac death. Upper airway collapse during sleep can induce ventricular electromechanical decoupling, creating an arrhythmogenic substrate (45). In an observational cohort study of 10,701 adults undergoing a diagnostic clinical polysomnogram, resuscitated or fatal sudden cardiac arrest was best predicted by an AHI ≥ 20 events/hour, mean nocturnal oxygen saturation < 93%, and lowest oxygen saturation ≤ 78% (14). Thus, SDB may increase the risk of sudden cardiac death in part through the effects of nocturnal intermittent hypoxemia. *In vivo* experimental studies show that chronic intermittent hypoxemia can augment sympathetic activity, alter ventricular repolarization, and increase the risk of sudden cardiac death (46). Hypoxia has been reported to increase the late component of the sodium current and alter the L-type calcium current *in vitro* (47–49). Further, hypoxia regulates the L-type calcium channels by increasing their sensitivity to β-adrenergic stimulation (50). Significant instability in action potential duration, including early afterdepolarizations, is detected when hypoxia is combined with activation of the sympathetic nervous system using isoproterenol in adult guinea pig myocytes or the Luo-Rudy *in silico* model (51). Therefore, the hypoxemia and the sympathetic activation associated with SDB are expected to increase repolarization lability.

QTVI was proposed by Berger et al. (37) as a measure of ventricular repolarization lability to forecast the occurrence of reentrant ventricular arrhythmias in patients with dilated cardiomyopathy. In the absence of cardiac pathology, QTVI increases with age and is higher in women than men (52). Studies that have characterized QTVI have consistently found that a larger QTVI is associated with a greater risk for ventricular arrhythmias, sudden cardiac death, and both all-cause and cardiovascular mortality (17–22). However, there have been major gaps in our understanding of the association between SDB and QTVI, and available data are limited and conflicting due to methodological differences in study design and the lack of control for confounding factors (29–31). Inconsistent relationships have been reported between the presence of SDB and the measures of QT variability that do not account for the effect of heart rate (53). QTVI is a dimensionless index that measures the beat-to-beat changes (as opposed to using a median/average signal) in QT that are disproportionate to the changes in heart rate. This normalization makes QTVI a more reliable marker of proarrhythmic risk, as it provides a more accurate assessment of repolarization lability that is less influenced by heart rate variations, which are common in SDB. The data presented herein provide support for the role of SDB in increasing QTVI, given the availability of cross-sectional, longitudinal, and experimental evidence. By minimizing confounding, the current study also implicates ventricular repolarization instability as a potential mediator of all-cause mortality in SDB.

The argument for a causal link between SDB, ventricular repolarization instability, and mortality is furthered by the observation in the current study that acute exposure to intermittent hypoxia for as little as four hours increased ventricular repolarization lability. The lack of an association between the frequency of arousals from sleep and QTVI in the current study could be related to the fact that arousal scoring in SHHS had poor reproducibility, which could dampen the ability to detect potential associations. Recurrent arousals from sleep increase sympathetic nervous system activity (54), which can increase ventricular repolarization lability and arrhythmogenicity. In fact, data from an epidemiological study of older men showed that excessive QTVI during arousals predicted all-cause and cardiovascular mortality (55). Such arousal-induced increases in QTVI appeared to be transient, as there were no associations between pre-arousal QTVI and mortality. Our results, however, suggest a cumulative impact of intermittent hypoxemia on cardiac repolarization. Indeed, in the longitudinal analysis, the mean QTVI increased to -0.86 in those with incident SDB without any concurrent medical conditions. The high level of ventricular repolarization lability present with incident SDB is comparable to that seen in heart failure patients with an increased risk for mortality (22). Additionally, the QTVI levels seen with incident SDB were similar to that of patients on chronic methadone therapy, who experience nocturnal hypoxemia and a predisposition for high density of premature ventricular contractions during sleep (56). Methadone, an opioid that inhibits both hERG and I_K1_ currents (57), is associated with a significant risk for arrhythmias and sudden nocturnal death (58). The present study suggests that a similar proarrhythmic effect might be associated with intermittent hypoxemia and sympathetic stimulation.

Strengths of the current study include minimizing the effects of confounding covariates by rigorous matching of study participants for the cross-sectional comparisons and assessing the effects of incident SDB on QTVI, and the use of full-montage polysomnography to characterize SDB and ventricular repolarization abnormalities during sleep. The experiment demonstrating an increase in QTVI under conditions of exposure to acute intermittent hypoxia holds significant relevance for causal inferences on the effects of SDB-related intermittent hypoxemia on QTVI. Limitations of the current study include the lack of ascertaining a dose-response association between SDB and QTVI, and using a community-based versus a clinical sample which may influence overall inferences regarding QTVI in SDB.

The current study motivates investments in future efforts to determine whether treatment of SDB with positive airway pressure therapy is associated with improved ventricular repolarization and downstream effects on mortality, particularly in at-risk patients such as those with congestive heart failure.

## Data Availability

Anonymized data of the first two cohorts are available from National Sleep Research Resource. Anonymized data of the third cohort are available from the corresponding author upon reasonable request.

## Disclaimer

The opinions and assertions expressed herein are those of the authors and do not reflect the official policy or position of the Uniformed Services University of the Health Sciences or the Department of Defense.

