## Supplemental Tables for "Sleep-Disordered Breathing Destabilizes Ventricular Repolarization"

**Table S1** compares matched participants with severe SDB and without SDB stratified by NREM and REM sleep. Mean $SpO_2$, minimum $SpO_2$, QTVI, heart rate, and SDNN were larger in participants with severe SDB than without SDB during NREM and REM sleep.

**Table S2** displays the effects of incident SDB on mean $SpO_2$, minimum $SpO_2$, QTVI, heart rate, and SDNN stratified by NREM and REM sleep.

**Table S3** summarizes the measures of oxygen saturation levels ($SpO_2$) under intermittent hypoxia and normoxia conditions.

**Table S1. Stratified analysis of electrocardiographic parameters and oxygen saturation levels for the cross-sectional sample of the SHHS participants with severe SDB versus those without SDB.**

| Parameters | No SDB | Severe SDB | *P* value |
|---|---|---|---|
| NREM | | | |
| Mean $SpO_2$ (%) | 94.98 (0.23) | 93.80 (0.20) | < 0.001 |
| Minimum $SpO_2$ (%) | 94.53 (0.23) | 91.53 (0.27) | < 0.001 |
| QTVI | -1.43 (0.07) | -1.19 (0.08) | 0.033 |
| HR (beats/min) | 65.03 (1.17) | 68.14 (1.02) | 0.048 |
| SDNN (msec) | 32.88 (2.36) | 42.12 (3.42) | 0.028 |
| REM | | | |
| Mean $SpO_2$ | 95.27 (0.26) | 93.22 (0.29) | < 0.001 |
| Minimum $SpO_2$ | 94.58 (0.28) | 90.00 (0.50) | < 0.001 |
| QTVI | -1.41 (0.06) | -1.15 (0.09) | 0.019 |
| HR | 65.69 (1.11) | 68.90 (1.22) | 0.053 |
| SDNN | 32.63 (2.31) | 40.62 (3.04) | 0.038 |

Values are the mean (SEM). *P* values are for the Student's *t* test comparing matched participants with and without SDB. N = 61 for each group.

**Table S2. Stratified analysis of electrocardiographic parameters and oxygen saturation levels for the longitudinal sample of the participants with and without incident SDB.**

| Parameters | Baseline | | *P* value | Follow-up | | *P* value |
|---|---|---|---|---|---|---|
| | No SDB | Incident SDB | | No SDB | Incident SDB | |
| NREM | | | | | | |
| Mean $SpO_2$ (%) | 95.75 (0.22) | 94.96 (0.24) | 0.018 | 95.54 (0.23) | 91.34 (0.15) | < 0.001 |
| QTVI | -1.28 (0.10) | -1.23 (0.15) | 0.778 | -1.24 (0.10) | -0.86 (0.14) | 0.031 |
| HR (beats/min) | 63.76 (1.50) | 64.63 (1.73) | 0.706 | 63.93 (1.75) | 66.87 (2.07) | 0.283 |
| SDNN (msec) | 29.71 (3.07) | 27.19 (2.48) | 0.526 | 27.31 (2.36) | 24.11 (3.28) | 0.434 |
| QTc (msec) | 436.27 (5.45) | 447.47 (8.50) | 0.273 | 444.66 (5.29) | 450.70 (6.13) | 0.459 |
| REM | | | | | | |
| Mean $SpO_2$ | 96.11 (0.24) | 95.15 (0.24) | 0.007 | 95.84 (0.26) | 90.51 (0.25) | < 0.001 |
| QTVI | -1.18 (0.10) | -1.31 (0.17) | 0.493 | -1.27 (0.10) | -1.01 (0.15) | 0.163 |
| HR | 65.35 (1.45) | 65.17 (1.96) | 0.943 | 65.75 (1.97) | 65.55 (2.20) | 0.945 |
| SDNN | 25.93 (2.14) | 30.54 (3.86) | 0.301 | 28.06 (2.94) | 25.99 (3.22) | 0.636 |
| QTc | 440.08 (5.83) | 447.43 (8.72) | 0.484 | 446.94 (5.91) | 445.69 (5.03) | 0.873 |

Values are the mean (SEM). *P* values are for the Student's *t* test comparing participants with and without incident SDB.

**Table S3. Oxygen saturation levels during normoxia and intermittent hypoxia.**

| $SpO_2$ (%) | Normoxia | Intermittent Hypoxia | *P* value |
|---|---|---|---|
| Mean | 97.37 (0.20) | 90.90 (0.20) | < 0.001 |
| Minimum | 96.11 (0.29) | 81.05 (0.50) | < 0.001 |
| SD | 0.56 (0.03) | 4.53 (0.14) | < 0.001 |

Values are the mean (SEM). *P* values are for the paired samples *t* test between normoxia and intermittent hypoxia experiments. N = 19.
